# Evaluation of Nowcasting for Real-Time COVID-19 Tracking — New York City, March–May 2020

**DOI:** 10.1101/2020.10.18.20209189

**Authors:** Sharon K. Greene, Sarah F. McGough, Gretchen M. Culp, Laura E. Graf, Marc Lipsitch, Nicolas A. Menzies, Rebecca Kahn

## Abstract

To account for delays between specimen collection and report, the New York City Department of Health and Mental Hygiene used a time-correlated Bayesian nowcasting approach to support real-time COVID-19 situational awareness. We retrospectively evaluated nowcasting performance for case counts among residents diagnosed during March–May 2020, a period when the median reporting delay was 2 days. Nowcasts with a 2-week moving window and a negative binomial distribution had lower mean absolute error, lower relative root mean square error, and higher 95% prediction interval coverage than nowcasts conducted with a 3-week moving window or with a Poisson distribution. Nowcasts conducted toward the end of the week outperformed nowcasts performed earlier in the week, given fewer patients diagnosed on weekends and lack of day-of-week adjustments. When estimating case counts for weekdays only, metrics were similar across days the nowcasts were conducted, with Mondays having the lowest mean absolute error, of 183 cases in the context of an average daily weekday case count of 2,914. Nowcasting ensured that recent decreases in observed case counts were not overinterpreted as true declines and supported health department leadership in anticipating the magnitude and timing of hospitalizations and deaths and allocating resources geographically.

## Introduction

Timeliness is a key attribute of surveillance systems for reportable infectious diseases (1, 2). Timely COVID-19 surveillance data are used by governments and communities to allocate resources and to decide when to tighten or loosen physical distancing and other prevention measures (3, 4). However, public health authorities track reportable diseases at a lag, given delays from infection to symptom onset, care seeking, specimen collection, laboratory testing, and reporting (5). Monitoring pre-diagnostic data sources (e.g., emergency department syndromic surveillance (6), internet searches and social media (7), participatory surveillance of self-reported symptoms (8), smart thermometers (9), etc.) can improve timeliness at the expense of specificity, such as an inability to distinguish increases in respiratory illness attributable to influenza from COVID-19. Another approach that preserves specificity when monitoring COVID-19 disease trends is to leverage partially reported disease data, formally accounting for data lags.

The terms “nowcasting,” or predicting the present, and “hindcasting,” or predicting through the day prior to the present, describe a wide range of statistical adjustments used to fill in cases that are not-yet-reported, offering health officials a more up-to-date picture for situational awareness (10). For example, researchers have assessed the potential to nowcast COVID-19 cases and deaths using Google Trends data available in near-real time (11), and have applied a range of modeling approaches that leverage reporting delays to estimate the number of not-yet-reported cases and deaths (12, 13). Using mathematical models to exploit COVID-19 transmission dynamics, nowcasting also has been extended to COVID-19 forecasting systems (14, 15). In a majority of these approaches, the nowcasting mechanism relies on accurately estimating the distribution of reporting delays; however, infectious disease transmission contains an important temporal component, in that incidence is correlated from one time point to the next, which has also been shown to be important in nowcasting (10). We describe the use and evaluation of a time-correlated Bayesian nowcasting approach at the New York City Department of Health and Mental Hygiene (NYC DOHMH) during the first epidemic wave of COVID-19 to support real-time situational awareness and resource allocation.

## Methods

### Reportable disease surveillance data

#### Persons tested

Clinical and commercial laboratories are required to report all results (including positive, negative, and indeterminate results) for SARS-CoV-2 tests for New York State residents to the New York State Electronic Clinical Laboratory Reporting System (ECLRS) (16, 17). For NYC residents, ECLRS transmits reports to NYC DOHMH. These laboratory reports include specimen collection date and patient demographic information, including residential address.

For nowcasting persons newly tested, NYC DOHMH deduplicated laboratory reports, retaining the first report received (“report date”) in ECLRS per person of a SARS-CoV-2 polymerase chain reaction (PCR) test. We retained the first specimen collection date for that associated test report date and the patient’s ZIP code of residence at time of report.

ZIP codes are collections of points constituting a mail delivery route. The United States Census Bureau developed ZIP code tabulation areas, which are aggregates of census blocks, to provide an areal representation of ZIP codes. NYC DOHMH created a custom geography referred to as modified ZCTA (modZCTA) by merging ZCTAs with populations <3000 to an adjacent ZCTA with a larger population and merging interior ZCTAs with smaller populations to the surrounding ZCTA (18, 19). There are 177 modZCTAs within NYC.

#### Confirmed cases

At NYC DOHMH, electronic laboratory reports are automatically standardized, and positive results indicating a confirmed case (i.e., detection of SARS-CoV-2 RNA in a clinical specimen using a molecular amplification detection test) (20) are transmitted to the NYC DOHMH’s communicable disease surveillance database (Maven, Conduent Public Health Solutions, Austin, Texas). For confirmed cases, the “diagnosis date” was defined as the specimen collection date of the first positive test. The “report date” was defined as the date the case was created in the disease surveillance database, which typically corresponded to the date the first positive test was reported to ECLRS.

Hospitalization status was ascertained by routinely matching demographic data for confirmed COVID-19 cases with hospitalized patients in supplemental data systems, including regional health information organizations, the New York State Hospital Emergency Response Data System, and NYC public hospitals (21). For each hospitalized patient with a confirmed COVID-19 diagnosis, the hospital name for the most recent hospitalization in NYC was standardized to the name of a fully operational medical center. Patients with hospital discharge dates >14 days prior to the collection date of their first positive PCR result were not considered hospitalized for COVID-19. The date of hospitalization ascertainment was not retained.

### Real-time nowcasting

NYC DOHMH nowcasted 3 outcomes (i.e., confirmed cases, ever-hospitalized cases, and persons tested) among NYC residents at weekly increments, on Mondays using reports received through the prior day on Sunday, in real-time through May 2020. Starting March 24, 2020, nowcasts were conducted for all confirmed COVID-19 cases and restricted to the subset of confirmed COVID-19 cases among patients ever hospitalized. Starting May 2, 2020 as testing became more widely available (22), nowcasts were conducted for persons newly tested by PCR for SARS-CoV-2. Each outcome was nowcasted citywide and also stratified by modZCTA of patient residence, to support targeting of community-based resources. Hospitalized cases were also nowcasted stratifying by health care facility, to support allocating resources to hospitals.

To account for reporting delays and the shape of the outcome-specific epidemic curve, we applied the R package Nowcasting by Bayesian Smoothing (NobBS) (10, 23) to data for specimens collected or diagnoses during the 3 weeks prior to the nowcast through the date prior to the nowcast, assuming a Poisson distribution. Briefly, this approach corrects for underestimation of cases in real-time caused by delays in reporting, learning the historical distribution of delays and relationship between cases in sequential time points to estimate the number of cases not-yet-reported. The 3-week moving window was selected to balance recency with stability. In performing stratified nowcasts, NobBS estimated the delay distribution citywide and the epidemic curve uniquely by stratum. Reports visualizing nowcast results were distributed weekly to DOHMH leadership for situational awareness.

### Retrospective nowcasting evaluation

For the outcome of confirmed COVID-19 cases, we assessed the sensitivity of nowcasting results for patients diagnosed citywide during March 22–May 31, 2020 (excluding cases diagnosed during March 1–21 given limited testing) to several choices: [1] day-of-week when the nowcast was performed, given outpatients with milder illness sought care and were diagnosed less frequently on weekends, when health care provider offices were typically closed or had more limited hours, [2] window length, given time-varying SARS-CoV-2 testing availability and uptake in NYC, and [3] assumed underlying distribution (Poisson or negative binomial) for case occurrence. In addition, for nowcasting the number of cases stratified by modZCTA, we compared results using [1] the “strata” option in NobBS — which estimated the delay distribution citywide and epidemic curve separately for each modZCTA — vs. estimating both the delay distribution and epidemic curve separately for each modZCTA, and [2] 10,000 vs. 3,000 adaptations when optimizing the nowcasting algorithm (10).

Data for the evaluation were frozen as of June 30, 2020, capturing reports received through 1-month after the end of the assessment period. We mimicked prospective surveillance at weekly intervals and daily temporal resolution, retaining the number of estimated cases for each of the prior 7 days (i.e. 1-7 day hindcasts). We used the mean absolute error and the average daily relative root mean square error across all days evaluated to compare the point estimate of the number of daily hindcasted cases over the time series to the true number of cases reported. For each of these metrics, lower numbers indicate better performance of the hindcast. We also assessed the 95% prediction interval coverage, i.e., the proportion of days during the study period when the 95% prediction interval included the true number of cases (10), which should ideally be 95%.

This work was reviewed and deemed public health surveillance that is non-research by the DOHMH Institutional Review Board.

## Results

Among confirmed COVID-19 cases residing in NYC and diagnosed during March–May 2020, the median delay between specimen collection and report was 2 days (interquartile range [IQR]: 1–4 days; 90^th^ percentile: 7 days). By month of report for diagnoses during March–May 2020, the median (IQR; 90^th^ percentile) number of days for this delay for reports received in March 2020 was 2 (1–4; 7), in April was also 2 (1–4; 7), in May was 2 (1–3; 5), and in June (given the study period included cases diagnosed through May) extended to 7 (4–19; 62). Hindcasts were performed weekly on Mondays in real-time, with results visualized for DOHMH leadership (e.g., Figure 1). However, the retrospective performance evaluation determined that real-time hindcasts on Mondays using a 3-week window and an assumed Poisson distribution more often overestimated than underestimated the number of not-yet-reported cases and resulted in overly narrow 95% prediction intervals (Figure 2 and and Web Figure 1).

**Figure 1.**
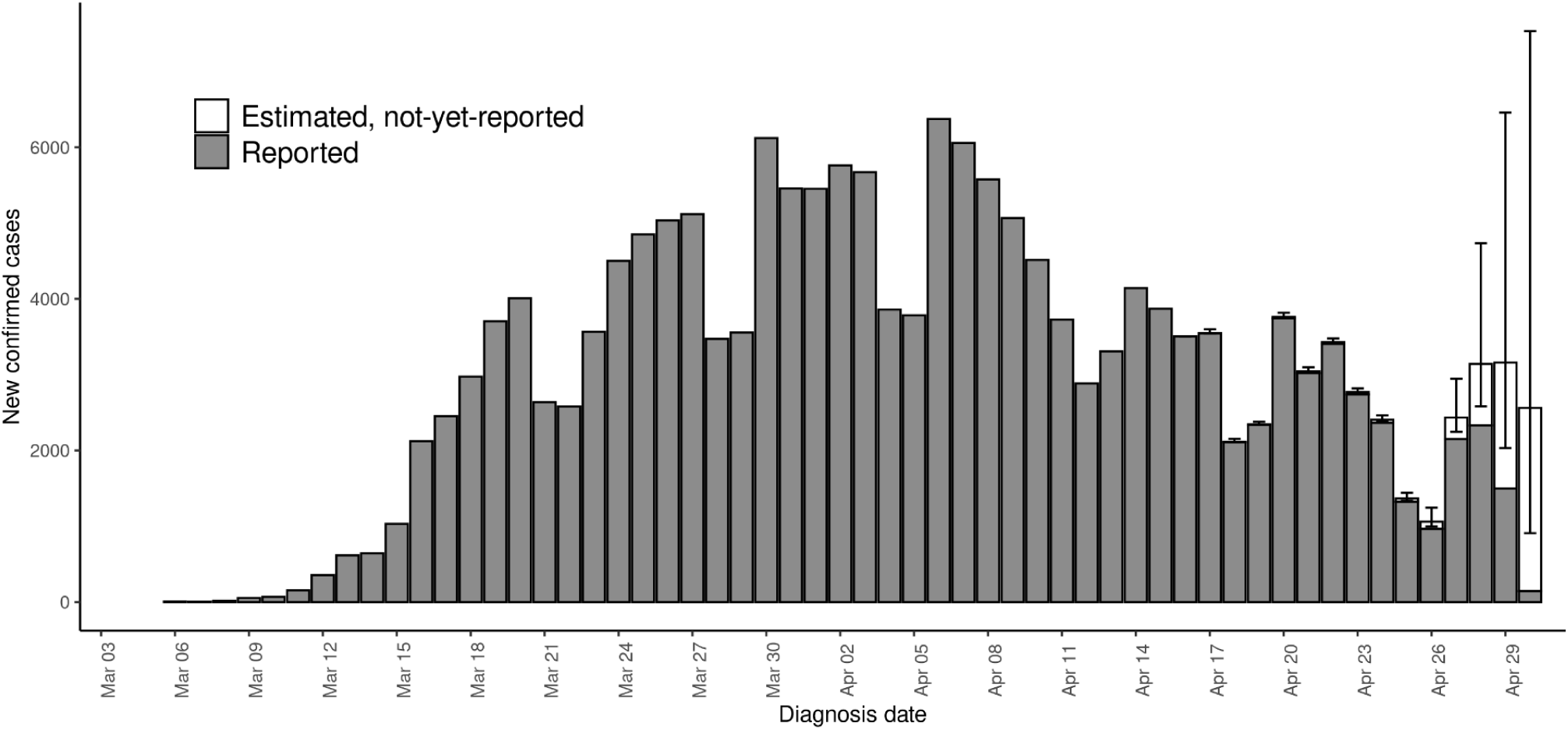
Example hindcast visualization of epidemic curve of reported and estimated but not-yet-reported number of confirmed cases among New York City residents diagnosed with COVID-19, March 1–April 30, 2020. Illustrative hindcast performed using cases reported through April 30, 2020 (i.e., a Thursday), a 2-week moving window, and a negative binomial distribution.

**Figure 2.**
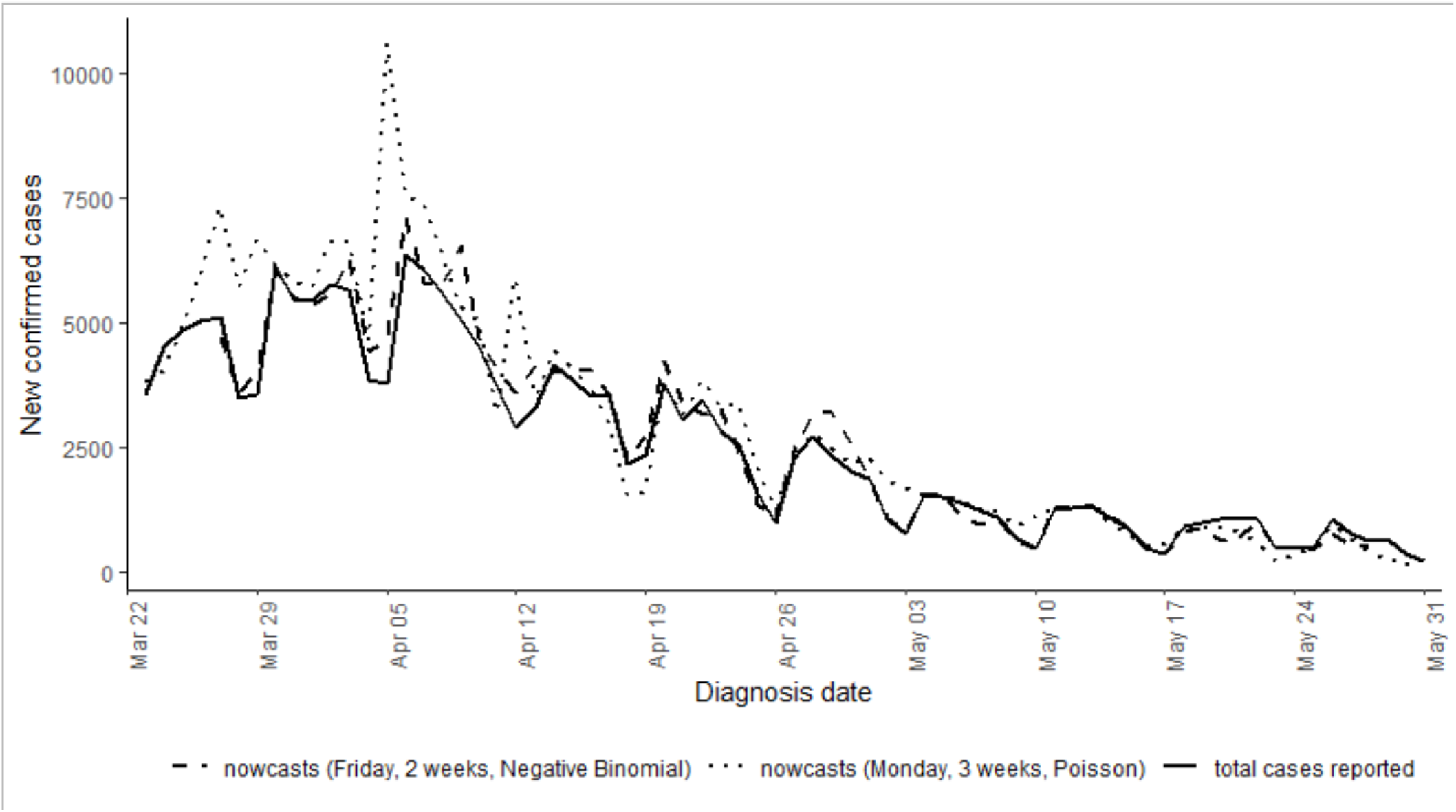
Comparison of 7-day hindcasts conducted on Fridays with a 2-week window and negative binomial distribution and 7-day hindcasts conducted on Mondays with a 3-week window and Poisson distribution. Total cases reported as of June 30, 2020 shown with black line.

We found that citywide hindcasts with a 2-week moving window and a negative binomial distribution had a 44% lower mean absolute error, a 31% lower relative root mean square error, and 0.65 higher 95% prediction interval coverage than hindcasts conducted with a 3-week moving window or with a Poisson distribution (Table 1, Web Table 1, Web Figures 1 & 2). Hindcasts conducted towards the end of the week (Thursday–Saturday) performed better than hindcasts performed earlier in the week, presumably as they had the furthest distance from the weekends. Weekends had lower overall case counts than weekdays (Figure 1). Until mid-May, hindcasts more often overestimated than underestimated true cases counts, whereas at the end of May hindcasts more often underestimated case counts, reflecting changes in the delay distribution over time (Figure 1, Web Figure 3).

**Table 1.** Performance Measures for Hindcasting Approaches Applied to Citywide Case Counts of New York City Residents Diagnosed with COVID-19, March 22–May 31, 2020. The first value in each column represents the metric across all days nowcasted, and the second assesses only weekdays.

| **Sensitivity analysis** | **Approach** | **Mean absolute error** | **Relative root mean square error** | **95% prediction interval coverage** |
| --- | --- | --- | --- | --- |
| Base scenario used in near real-time by NYC DOHMH, using 3-week window with Poisson distribution | All days | 544; 559 | 0.20; 0.19 | 0.16; 0.16 |
|  | Hindcasting each Monday for the previous Monday–Sunday | 556, 338 | 0.25, 0.12 | 0.14; 0.20 |
| Day-of-week hindcasting was performed for previous 7-day period (2-week window, negative binomial distribution) | All days | 306; 258 | 0.14; 0.10 | 0.81; 0.84 |
|  | Monday | 336, 183 | 0.20, 0.07 | 0.86, 0.82 |
|  | Tuesday | 335, 233 | 0.16, 0.08 | 0.83, 0.84 |
|  | Wednesday | 307, 275 | 0.14, 0.11 | 0.81, 0.87 |
|  | Thursday | 271, 257 | 0.11, 0.11 | 0.81, 0.84 |
|  | Friday | 255, 267 | 0.10, 0.11 | 0.75, 0.84 |
|  | Saturday | 260, 267 | 0.11, 0.11 | 0.73, 0.80 |
|  | Sunday | 372, 273 | 0.16, 0.10 | 0.87, 0.88 |

To minimize day-of-week effects that were most prominent on weekends, we also restricted performance analysis to hindcasts of cases on weekdays only, which resulted in better metrics, as expected (Table 1 and Web Table 1). The hindcasts restricted to estimating case counts for weekdays with a 2-week moving window and negative biniomial distribution also performed better than the hindcasts with a 3-week moving window and Poisson distribution, with 54% lower mean absolute error, 46% lower relative root mean square error and 0.69 higher 95% prediction interval coverage (Table 1, Web Table 1). Performance metrics were similar across days the hindcasts were conducted, with Mondays having the lowest mean average error and relative root mean square error, as expected given the 2 additional days between the last day reported (Friday) and the day the hindcast was conducted (Monday). On weekdays during the study period, the average daily case count after data lags resolved was 2,914, the average hindcasted case count with a 2-week window and negative binomial distribution conducted on Mondays was 2,878, and the mean absolute error was 183.

For hindcasts at the modZCTA-level, a 2-week moving window and negative binomial distribution performed best across all metrics evaluated (Table 2 and Web Table 1), although the prediction interval coverage for the nowcasts with a Poisson distribution was higher than for citywide hindcasts. The hindcasts that assumed a citywide delay distribution performed slightly better than hindcasts that assumed different distributions by modZCTA. Metrics for 3,000 vs. 10,000 adapations were essentially the same.

**Table 2.** Performance Measures for Hindcasting Approaches Applied to Case Counts of New York City Residents Diagnosed with COVID-19, March 22–May 31, 2020, Stratified by Modified ZIP Code Tabulation Area of Residence (modZCTA). The first value in each column represents the metric across all days hindcasted, and the second assesses only weekdays.

| **Sensitivity analysis** | **Approach** |  | **Mean absolute error** | **Relative root mean square error** | **95% prediction interval coverage** |
| --- | --- | --- | --- | --- | --- |
| Base scenario used in near real-time by NYC DOHMH | “strata” option in NobBS (which estimated the delay distribution citywide and epidemic curve separately for each modZCTA), conducted on Mondays | 3-week Poisson (10,000 nAdapt) | 3.82; 2.75 | 0.37; 0.18 | 0.84; 0.84 |
|  |  | 3-week Poisson (3,000 nAdapt) | 3.83; 2.76 | 0.37; 0.18 | 0.84; 0.84 |
|  |  | 2-week negative binomial (10,000 nAdapt) | 2.92; 2.09 | 0.33; 0.15 | 0.93; 0.93 |
|  |  | 2-week negative binomial (3,000 nAdapt) | 2.93; 2.08 | 0.34; 0.15 | 0.93; 0.93 |
| Conducting hindcasts on Fridays | “strata” option in NobBS (which estimated the delay distribution citywide and epidemic curve separately for each modZCTA), conducted on Fridays | 2-week negative binomial | 2.62; 2.98 | 0.22; 0.25 | 0.94; 0.95 |
| Estimate delay distribution separately by modZCTA | Estimating both the delay distribution and epidemic curve separately for each modZCTA, conducted on Mondays | 2-week negative binomial | 3.55; 2.57 | 0.36; 0.21 | 0.94; 0.95 |

## Discussion

NYC DOHMH improved situational awareness of COVID-19 testing and cases during the first epidemic wave in near-real time by applying NobBS, a readily accessible nowcasting and hindcasting method. As a result of the retrospective performance evaluation, to improve nowcast accuracy prospectively effective August 2020, we implemented the following changes to the nowcasting approach: (i) used a negative binomial case distribution instead of a Poisson, (ii) linked the determination of the moving-window length (2 or 3 weeks) to the 90^th^ percentile of the lag between specimen collection and report for reports received in the most recent week, choosing 3 weeks if the 90^th^ percentile of the lag distribution is >14 days, and (iii) suppressed nowcasting results for specimens collected on weekends, given lack of adjustment for day-of-week effects. The evaluation supported the results of nowcasting conducted on any weekday. A previous evaluation of influenza nowcasts also found a negative binomial distribution more appropriately captured uncertainty in case counts than a Poisson distribution (10).

Despite a mature electronic laboratory reporting system and strong informatics infrastructure and data cleaning procedures at NYC DOHMH, input data available for nowcasting had several limitations. First, for records with long lags between specimen collection and report, as long as the specimen was reported to have been collected during the pandemic period, it was not possible to distinguish long lags attributable to true delays in testing or reporting — and thus informative to the delay distribution — from long lags attributable to laboratory data entry errors in specimen collection dates. Second, nowcasting by patient modZCTA of residence relied on accurate laboratory reporting of patient address. For example, one week of real-time nowcasting results were biased when, for a batch of reports, one commercial laboratory misreported its own address as the residential address of all patients tested. Third, a large proportion of records had missing onset date. NobBS is designed for use with complete linelists with no missing onset or report dates. Given the complexities of imputing onset date from diagnosis date, nowcasts were instead conducted by specimen collection or diagnosis date. Fourth, patient hospitalization status was largely ascertained by matching administrative records. To allow time for record matching, hospitalization nowcasts were conducted at a 3-day lag, limiting the real time availability of results. Furthermore, records from certain facilities were unavailable in near-real time, so nowcasts of hospitalizations by patient residence and by facility were subject to spatial bias, although still considered by DOHMH leadership to be useful for situational awareness.

This version of NobBS (0.1.0) also had several limitations when applied for nowcasting COVID-19 in NYC. First, citywide nowcasting was based on a linelist with only 2 data elements: specimen collection or diagnosis date and report date. Stratified nowcasts included a third data element: modZCTA or health care facility. There was no built-in functionality in NobBS to account for additional observable factors influencing data lags, including day-of-week and holiday effects in outpatient testing, and time-varying testing backlogs at specific laboratories differentially processing specimens for residents across neighborhoods. Similarly, there was no functionality to account for temporal trends in testing, e.g., the time-varying ratio of number of tests performed to number of cases detected. Increased testing uptake strongly influenced the shape of the observed epidemic curve during the study period as testing criteria at public health laboratories were relaxed, commercial and hospital labs developed testing capacity, and additional testing sites were opened and promoted. Third, while 95% prediction intervals reflected uncertainty in the nowcasts themselves — encompassing uncertainty in the estimation of the delay distribution as well as in the time evolution of the epidemic curve — they did not reflect uncertainty introduced by the user-specified window length. In other words, the moving window length had the potential to change nowcast estimates considerably. Analysts found it challenging to select the optimal window length in real time, and, given competing priorities during a pandemic, busy DOHMH officials would not have had adequate time to consider multiple nowcast versions with different window lengths as sensitivity analyses. The retrospective analysis, which covered a period when the 90^th^ percentile of delay between specimen collection and report was 7 days, found that a 2-week nowcasting window resulted in higher performance than a 3-week window. This finding might not be generalizable to periods with more extensive reporting delays (24). Fourth, in generating geographically stratified nowcasts, the “strata” option in NobBS estimated the delay distribution citywide and epidemic curve separately for each modZCTA or health care facility stratum. For a highly transmissible infectious disease, nowcasting performance might be improved by considering spatial relationships across geographic strata, including spatial autocorrelation. Fifth, early in the pandemic when case counts were sparse, cumulative case counts with 95% prediction intervals were of interest to DOHMH leadership, but this functionality was not built into NobBS and required a separate solution, though is now in a development version of the package available at https://github.com/sarahhbellum/NobBS. Finally, although government officials have demonstrated interest in publicizing test percent positivity by report date (25, 26), which can be biased by data lags, NobBS did not have functionality to nowcast percentages as an outcome. NobBS could be used to separately nowcast persons testing positive and negative and then to calculate test percent positivity, but there is no functionality to appropriately account for the separate uncertainties in the numerator and denominator of this percentage.

## Conclusion

When tracking ongoing outbreaks using epidemic curves, public health officials recognize that data for recent days are incomplete because of reporting delays. Data lags can make it difficult for policymakers to discern in near real-time whether apparent decreases in recent case counts are the result of public health interventions, such as social distancing guidelines.

NYC DOHMH filled in COVID-19 epidemic curves using NobBS, which helped ensure that recent decreases in observed case counts were not overinterpreted as true declines in disease and supported the continuation of policies to reduce transmission. Nowcasting citywide case counts supported situational awareness and assisted DOHMH leadership in anticipating the magnitude and timing of hospitalizations and deaths. Nowcasting hospitalizations by health care facility was useful in helping to route patient transports and avoid overburdening facilities. As the COVID-19 pandemic continues and jurisdictions brace for second waves of infections, state and local health departments should incorporate nowcasting into their workflows.

## Data Availability

Line-level data, as required for nowcasting using the NobBS R package, are not publicly available in accordance with patient confidentiality and privacy laws. Publicly available data are linked below.

https://www1.nyc.gov/site/doh/covid/covid-19-data.page

## Acknowledgments

The authors thank the NYC DOHMH Incident Command System Surveillance and Epidemiology Section, including Jennifer Baumgartner, Eric R. Peterson, and Miranda S. Moore for data preparation, Samia Baig for visualization, and Dr. Annie D. Fine for proposing nowcasting by health care facility. The authors also thank Angel Aponte for administering the DOHMH R server.

S.K.G. was supported by the Public Health Emergency Preparedness Cooperative Agreement (grant NU90TP922035-01), funded by the U.S. Centers for Disease Control and Prevention. R.K. was supported by Award Number U54GM088558 from the U.S. National Institute of General Medical Sciences. M.L. was supported by the Morris-Singer Fund and a subcontract from Carnegie Mellon University under U.S. Centers for Disease Control and Prevention Award U01IP001121.

This article’s contents are solely the responsibility of the authors and do not necessarily represent the official views of the Centers for Disease Control and Prevention, the National Institutes of Health, or the Department of Health and Human Services.

---

**Web Table 1.**
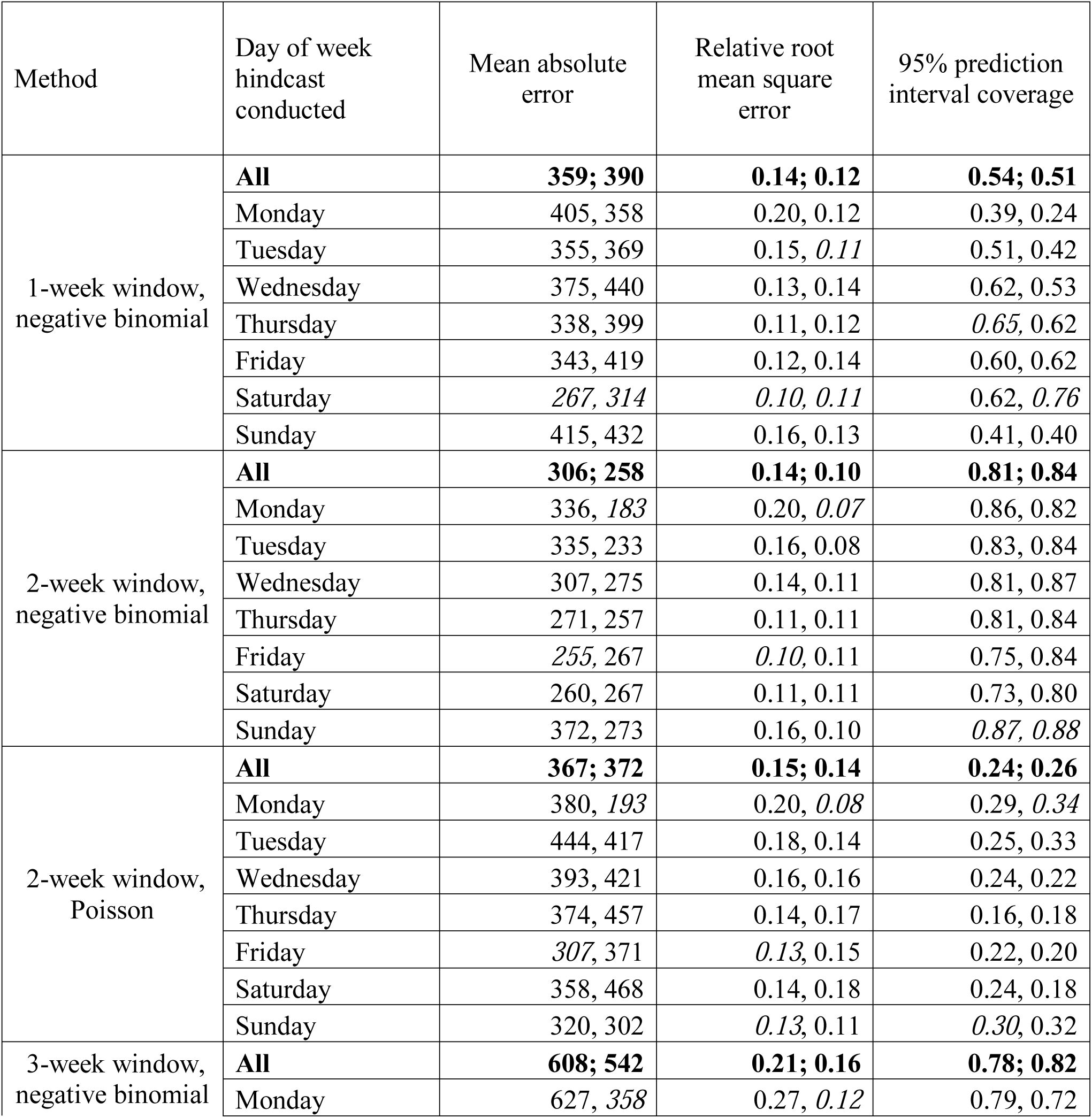
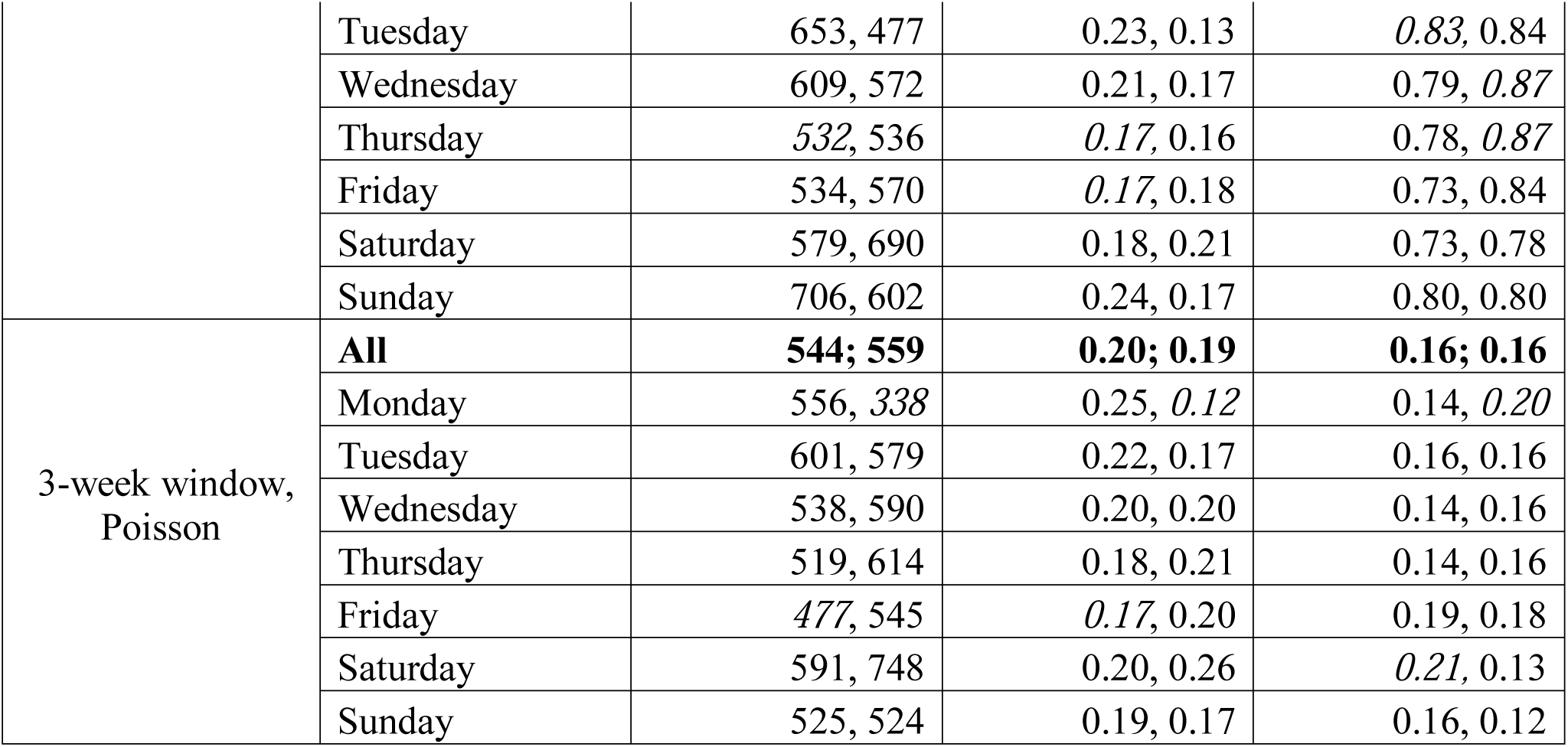
Performance measures for additional hindcasting approaches applied to citywide case counts of New York City residents diagnosed with COVID-19, March 22–May 31, 2020. The first value in each cell represents the metric across all days hindcasted, and the second assesses only weekdays. For each method and metric, the best performing day of week is italicized.

**Web Figure 1.**
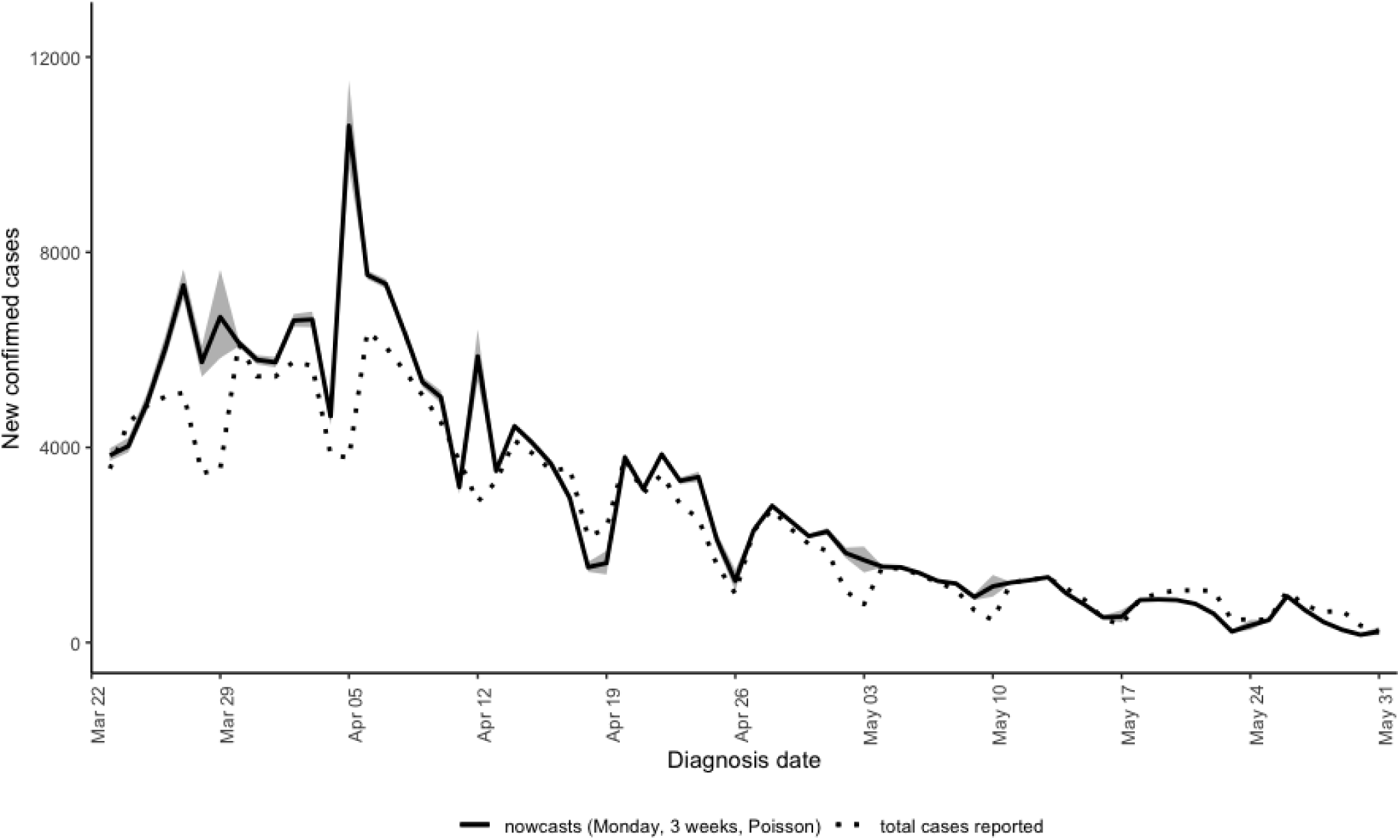
Estimates (black) with 95% confidence bounds (grey) for hindcasts conducted on Mondays, with a 3-week window and Poisson distribution.

**Web Figure 2.**
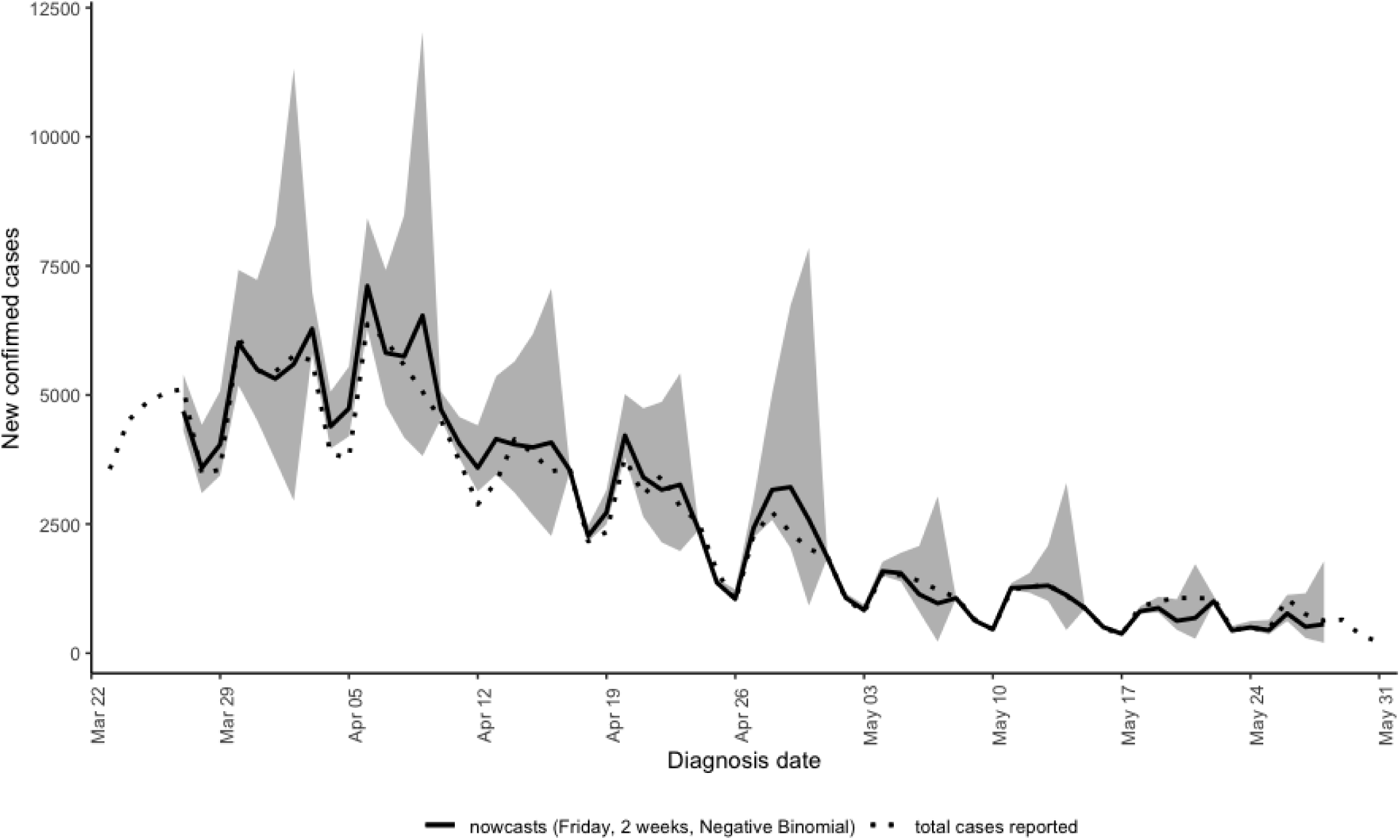
Estimates (black) with 95% confidence bounds (grey) for hindcasts conducted on Fridays, with a 2-week window and negative binomial distribution.

**Web Figure 3.**
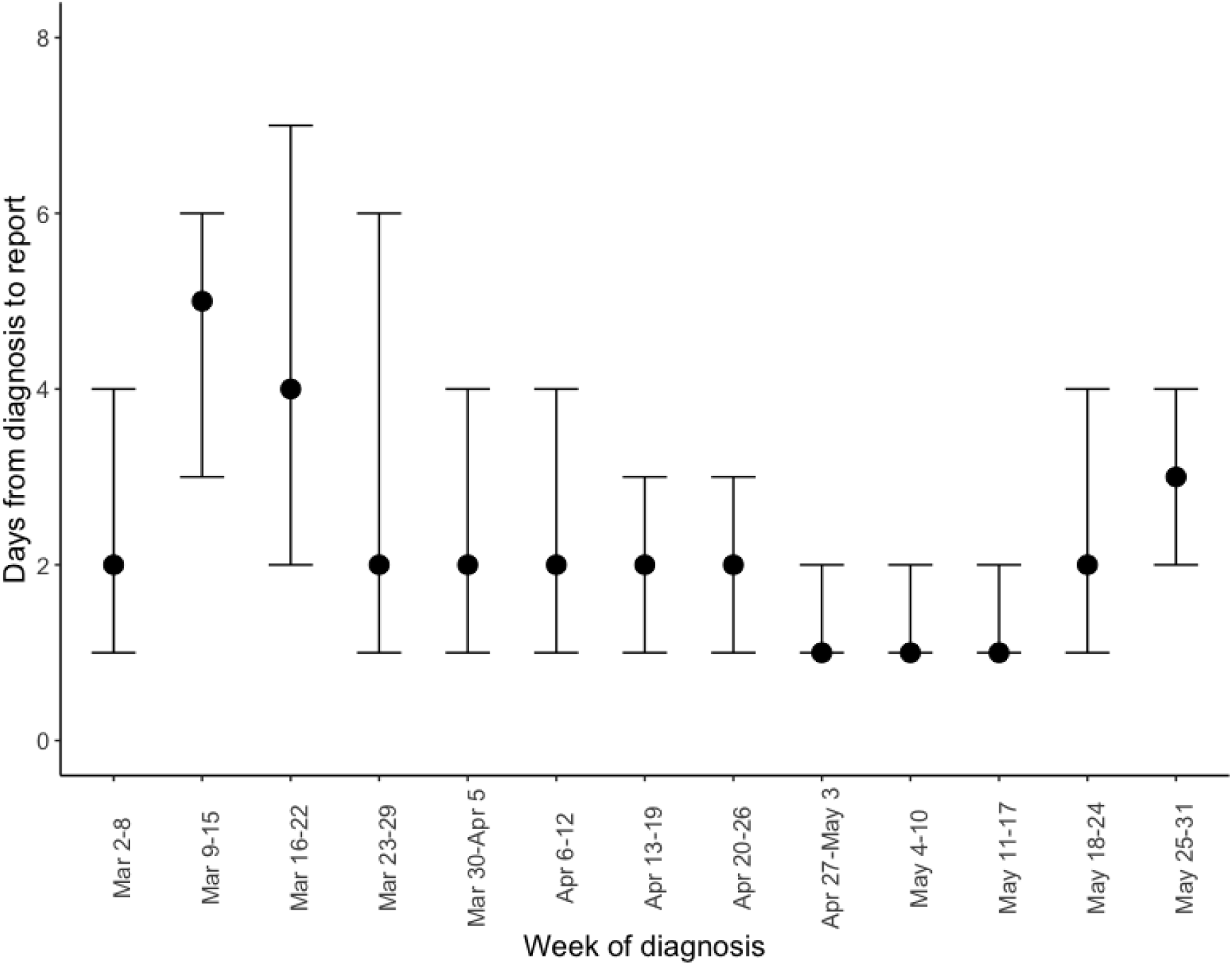
Median (interquartile range) of delays by week.

